# Data leakage in deep learning studies of translational EEG

**DOI:** 10.1101/2024.01.16.24301366

**Authors:** Geoffrey Brookshire, Jake Kasper, Nicholas Blauch, Yunan “Charles” Wu, Ryan Glatt, David A. Merrill, Spencer Gerrol, Keith J. Yoder, Colin Quirk, Ché Lucero

**Author notes:** Equal contributions (Geoffrey Brookshire, Jake Kasper).

## Abstract

A growing number of studies use deep neural networks (DNNs) to identify diseases from recordings of brain activity. DNN studies of electroencephalography (EEG) typically use cross-validation to test how accurately a model can predict the disease state of held-out test data. In these studies, segments of EEG data are often randomly assigned to the training or test sets. As a consequence, data from individual subjects appears in both training and test data. Could high test-set accuracy reflect leakage from subject-specific representations, rather than patterns that identify a disease? We address this question by testing the performance of DNN classifiers using segment-based holdout (where EEG segments from one subject can appear in both the training and test sets), and comparing this to their performance using subject-based holdout (where individual subjects’ data appears exclusively in either the training set or the test set). We compare segment-based and subject-based holdout in two EEG datasets: one classifying Alzheimer’s disease, and the other classifying epileptic seizures. In both datasets, we find that performance on previously-unseen subjects is strongly overestimated when models are trained using segment-based holdout. Next, we survey the literature and find that the majority of translational DNN-EEG studies use segment-based holdout, and therefore overestimate model performance on new subjects. In a hospital or doctor’s office, clinicians need to diagnose new patients whose data was not used in training the model; segment-based holdout, therefore, does not reflect the real-world performance of a translational DNN model. When evaluating how DNNs could be used for medical diagnosis, models must be tested on subjects whose data was not included in the training set.

## Introduction

Translational neuroscience studies increasingly turn to deep neural network (DNN) models to find structure in neural data. The power of DNN models comes from their ability to discover patterns in the data that researchers would not have been able to specify. In this literature, DNNs have been trained on a variety of imaging techniques to identify a wide range of clinical conditions. Many of these studies use DNNs to diagnose diseases based on anatomical neuroimaging. For example, DNN models can identify Alzheimer’s disease (AD) using structural magnetic resonance imaging (MRI) ^74^, and a variety of cancers and brain injuries using CT scans ^29,35^. In addition to anatomical data, a large number of studies have used DNNs to identify diseases from functional neuroimaging data. For example, DNNs with functional MRI show promise for identifying AD, Autism spectrum disorders, attention-deficit/hyperactivity disorder (ADHD), and schizophrenia ^73^. Furthermore, DNNs have been used with electroencephalography (EEG) to study a variety of different neural and cognitive disorders ^17^.

Deep learning helps to reveal previously-unknown patterns in neuroimaging data, but it also presents researchers with subtle pitfalls. One set of challenges concerns how the data are split into separate training and test sets. The training set is used to fit the model’s parameters, and the test set is used to estimate the model’s performance on new data. (A third subset of the data is often held aside as a validation set, used to tune the model’s hyperparameters and to determine when to stop training the model.) In some cases, researchers train their model on one subset of the available data, and then evaluate the model’s performance on a separate test set. In other cases, researchers use cross-validation (CV) to train and test models on multiple subsets of the data. Under both of these approaches, researchers must be careful to avoid “data leakage” when splitting the data into training and test sets. Data leakage, which arises when information about the test set is present in the training set, results in a positively-biased estimate of the model’s performance ^36^. For example, in a data-mining competition focused on identifying patients with breast cancer, one team of researchers found that the patient ID number carried predictive information about cancer risk ^59^. These ID numbers may have appeared after compiling data from different medical institutions. Because the ID number was assigned based on patients’ diagnosis, it constitutes a source of data leakage ^59^. In general, data leakage occurs when an experimenter handles the data in a way that artificially introduces correlations between the training and test sets.

DNN models typically require a large amount of training data to perform well, but neural datasets are usually expensive and difficult to obtain. To increase the number of observations available to train the model, these studies often split a single neural recording into multiple samples, and use each sample as a separate observation during training or testing. For example, a 3D structural MR volume could be split into multiple 2D slices, and an fMRI time-series could be split into multiple segments of time ^74^. When multiple observations from a single subject are included in both the training and test sets, it constitutes data leakage: Instead of learning a generalizable pattern, these models could learn characteristics of the individual subjects in the training set, and then simply recognize those familiar subjects in the test set. As a result, these models perform well in the study’s test set, leading the researchers to believe they have a robust classifier. In new subjects, however, the model may fail to generalize. In fact, leakage of subject information does occur in a number of published MRI studies ^74^. Furthermore, leakage of subject-specific information is widespread in translational studies using optical coherence tomography (OCT), and leads to strongly inflated estimates of test accuracy ^65^.

Studies using DNNs with EEG are particularly susceptible to data leakage. In these studies, each subject’s full EEG time-series (lasting several minutes) is commonly divided up into brief segments (lasting several seconds) ^17^. Each segment is then used as a separate observation during training or testing. This segmentation procedure is meant to ensure that DNN models have enough training data to learn robust representations of the patterns that characterize a disease, and to prepare the data for commonly-used model architectures. However, EEG segmentation leads to data leakage if the same subjects appear in both the training and test sets. Segments of EEG from one subject are more similar to each other than to segments from different subjects ^18^. Instead of learning an abstract representation that would generalize to new subjects, a DNN model could therefore achieve high classification accuracy by associating a label with each subject’s idiosyncratic pattern of brain activity. As a consequence, randomly splitting EEG segments into training and test sets results in data leakage, and a biased estimate of test performance: accuracy is high on the researchers’ test set, but the classifier will generalize poorly to new subjects. In a clinical setting, this leads to an apparently-promising diagnostic tool that fails when applied to new patients. To avoid this kind of data leakage, all segments from a given subject must be assigned to only a single partition of the data (i.e. train *or* validation *or* test).

How does leakage of subject-specific information bias the results of translational DNN-EEG studies? Here we address this question by examining the effects of data leakage in two case studies, and then reviewing the published literature to gauge the prevalence of this leakage. In the case studies, we reproduce two convolutional neural network (CNN) architectures used by published studies – both of which used a train-test split that introduced data leakage. First, we use a CNN to classify subjects as either healthy or as having dementia due to Alzheimer’s disease. Second, we use a CNN to classify whether segments of time contain an epileptic seizure. In both datasets, we find that real-world performance is dramatically overestimated when data from individual subjects is included in both the training and test sets. In the literature review, we find that the majority of translational DNN-EEG studies suffer from data leakage due to data from individual subjects appearing in both the training and test sets.

## Methods

### Deep neural network analysis overview

To investigate how segment-based holdout leads to data leakage, we reproduced the model architectures from two published studies ^56,58^. The goal of these analyses was not to develop an optimal architecture, but rather to evaluate the impact of different cross-validation choices on the estimated model performance. We therefore re-used the published architectures and data processing pipelines without modification.

### Experiment 1: Alzheimer’s disease diagnosis

#### EEG data

EEG recordings were provided by the Pacific Neuroscience Institute ^25^. All procedures were approved by the St. John’s Cancer Institute Institutional Review Board (Protocol JWCI-19-1101) in accordance with the Helsinki Declaration of 1975. Patients were evaluated by a dementia specialist as part of their visit to a specialty memory clinic (Pacific Brain Health Center in Santa Monica, CA) for memory complaints. This evaluations included behavioral testing as well as EEG recordings. After these evaluations, subjects were selected by retrospectively reviewing charts for patients aged 55 and older seen between July 2018 and February 2021.

Patients received a consensus diagnosis from a panel of board-certified dementia specialists. Diagnoses were performed using standard clinical methods on the basis of neurological examinations, cognitive testing (MMSE ^23^ or MoCA ^54^), clinical history (e.g. hypertension, diabetes, head injury, depression), and laboratory results (e.g. vitamin B-12 levels, thyroid stimulating hormone levels, and rapid plasma regain testing). These tests were used to rule out reversible causes of memory loss and to diagnose SCI, MCI, and dementia. EEG data was not included in the diagnostic process. Cognitive impairment was diagnosed on the basis of MMSE (or MoCA scores converted to MMSE ^8^), with MCI diagnosed according to established criteria ^42^. MCI was distinguished from dementia on the basis of preserved independence in functional abilities, and a lack of significant impairment in social or occupational functioning. SCI was diagnosed in patients with subjective complaints but without evidence of MCI. Diagnostic categorization was based on the clinical syndromes ^42^, and did not consider disease etiology or subtypes within each stage.

EEG data were recorded at 250 Hz using the eVox System (Evoke Neuroscience), with a cap that included 19 electrodes following the International 10-20 system (FP1, FP2, F7, F3, Fz, F4, F8, T7, C3, Cz, C4, T8, P7, P3, Pz, P4, P8, O1, and O2). The full EEG session included a 5-minute block of eyes-open rest, a 5-minute block of eyes-closed rest, and a 15-minute go/no-go task. In this study, we analyzed only the eyes-open resting-state data. Recordings were low-pass filtered below 125 Hz, and split into non-overlapping segments of 2 s (500 samples) for model training. Channels were stacked to produce matrices of shape (500, 19) as model inputs.

We selected all 49 subjects in the dataset who were diagnosed with dementia due to Alzheimer’s disease (18 male, 31 female; age 73.9 *±* 6.8 years). As a comparison, we selected an equal number of subjects with subjective cognitive impairment (SCI; *n* = 49, 18 male, 31 female; age 63.9 *±* 11.4 years).

#### Architecture

Based on previous work ^56^, a basic 1D convolutional neural network was used to classify segments of time-series data as SCI or AD. Because our goal was to evaluate the effects of different cross-validation strategies, we re-used the published architecture without modification. This model learns temporal filters that are applied equivalently across each EEG channel. Progressing through the network, subsequent layers build more complex features that take into account a larger temporal receptive field, and some invariance is achieved through pooling over time. The model consisted of 4 convolutional layers, each followed by rectification, max pooling, and batch normalization; convolutional layers were followed by 2 dense fully-connected layers of 20 and 10 hidden units, respectively, each rectified, and finally a dense connectivity to the output layer with 2 units representing AD yes/no probability logits. All deep learning models were trained with Keras and Tensorflow. The exact Keras code used to specify the architecture can be found in the Appendix.

#### Training

Models were trained for 70 epochs without any early stopping or hyperparameter tuning. A batch size of 32, initial learning rate of 0.0001, and the Adam optimizer were used to optimize models. Training accuracy was computed and stored online during each epoch, and averaged across batches to report the training accuracy for each epoch. To visualize how quickly the models reached their final performance, test set accuracy was also computed after each epoch, averaged across batches. Since we reused the model architecture from prior published work, no model selection was performed; performing ongoing validation on the test is therefore not a source of data leakage. For segment-based holdout, data were split using 10-fold cross-validation (see ‘Cross-validation’ for details).

### Experiment 2: seizure detection

#### EEG data

We analyzed data from the Siena Scalp EEG Database ^19,20^ hosted on PhysioNet ^28^. These recordings were collected in accordance with the Declaration of Helsinki, and approved by the Ethical Committee of the University of Siena. Participants provided written informed consent before beginning data collection. This dataset includes recordings from 14 epilepsy patients (age 20-71 years, 9 male) digitized at 512 Hz with electrodes arranged following the International 10-20 system. Seizures in the data were labeled by an expert clinician. This dataset contains 47 seizures in approximately 128 hours of recorded EEG. To ensure that the data were balanced between seizure and non-seizure epochs, we selected non-seizure data from the beginning of each subject’s recordings to match the duration of their seizure-labeled data. This led to 47 min 21 s of data in each condition (1:34:42 s in total).

In contrast to the previous section where raw time series were used, EEG data were prepared for the classifier analysis in the frequency domain, following the approach used by Rashed-Al-Mahfouz and colleagues ^58^. Spectrograms were computed with a window length of 256 samples (0.5 s) overlapping by 128 samples (0.25 s), using a Hann taper. Spectrograms were then divided into segments of 1.5 s. As in the original study, we used the RGB representation of the spectrogram (viridis color-map), and exported as 224 x 224 x 3 images for training and testing with the CNN models.

#### Architecture

The goal of the simulations was primarily to evaluate the impact of different cross-validation choices, not to evaluate the architecture. Thus, the architecture of Rashed-Al-Mahfouz and colleagues ^58^ was used without modification. To handle 3D spectrogram data (vs. 2D time-series used in the previous section), a 2D convolutional neural network was used. This model learns 2D spectrotemporal features that are applied equivalently across the spectrogram. The model contains 4 convolutional layers, each followed by rectification, pooling, and batch normalization, followed by 2 hidden fully-connected layers of 256 and 512 units each, dropout, and a final classification layer of 2 units corresponding to seizure yes/no. The exact Keras code used to specify the architecture can be found in the Appendix.

#### Training

Models were trained for 70 epochs with no early stopping. We used the RMSProp optimimzer with a batch size of 32 and a learning rate of 0.00001. Training accuracy was computed and stored online during each epoch, and averaged across batches to report the training accuracy for each epoch. To visualize how quickly the models reached their final performance, test set accuracy was also computed after each epoch, averaged across batches. Since we reused the model architecture from prior published work, no model selection was performed; performing ongoing validation on the test is therefore not a source of data leakage.

#### Cross-validation

This study is primarily concerned with the consequences of different approaches to splitting the data between training and test sets. We assess two types of train-test split: (1) holding out individual segments of EEG data without regard for subject ID (“segment-based holdout”), and (2) holding out entire subjects, ensuring that all segments for a given subject appear in only the training or the test set (“subject-based holdout”).

#### Segment-based holdout

Segment-based cross-validation considers all EEG segments to be equivalent, and divides them into training and validation partitions without considering subject ID. This segment-holdout approach will lead to data leakage if there is statistical non-independence due to multiple EEG segments coming from each subject. Given *n* segments and *m* time-points per segments, we construct a matrix *X* of EEG segments of size (*n, m*), and a vector ***y*** of diagnostic label of length *n*. The cross-validation is a simple partition of the index vector ***α*** = *{*1, 2, …, *n};* into disjoint subsets ***α***_train_ and ***α***_test_. Where *X*_*i*_ gives the *i*th segments of *X*, we then have *X*_train_ = *{X*_*i*_*};∀i ∈* ***α***_train_, *X*_*val*_ = *{X*_*i*_*};∀i ∈* ***α***_test_, and ***y***_train_ = *{****y***_*i*_*};∀i ∈* ***α***_train_, ***y***_test_ = *{****y***_*i*_*};∀i ∈* ***α***_test_.

#### Subject-based holdout

Subject-based cross-validation takes into account which subject each EEG segment comes from. This approach enforces that each subject appears in only one partition of the cross-validation, ensuring there is no leakage of subject-level information across training and test sets. To create this split, we consider an additional subject vector ***s***, which is used to constrain the partition of *X* and ***y***. Concretely, rather than partitioning the index vector ***α***, we partition the unique subject vector ***s***_*u*_, which gives the unique entries of ***s***, and collect all corresponding segments from each subject contained in train and validation partitions into *α*_train_ and *α*_test_. This enforces the constraint that *s*_*i*_≠*s*_*j*_∀*i* ∈ ***α***_train_, *j ∈* ***α***_test_. To perform k-fold cross-validation, we first divide ***s***_*u*_ into *k* non-overlapping chunks, and each chunk to serve as the validation data in each fold of cross-validation, where the remaining *k −* 1 chunks are reserved for training.

#### Literature review

We searched the literature for studies that used deep learning with segments of EEG to classify a variety of diseases. We searched Google Scholar for papers investigating Alzheimer’s disease, Parkinson’s disease, attention-deficit/hyperactivity disorder (ADHD), depression, schizophrenia, and seizures. We then searched the references of these papers to identify additional appropriate publications for inclusion. This non-exhaustive search included 63 papers, all of which were published since 2018 and used deep learning to study one of the conditions named above.

Next, we examined how the training and test sets were determined in these studies. If a paper specified that the EEG recordings were split into segments, but did not specify that they used subjects as an organizing factor of the train-test split, we labeled that study as using ‘segment-based’ holdout. Some papers specifically stated that segments from individual subjects were included in both the training and test sets (for example, studies that trained separate models for each subject); these studies were also labeled as segment-based holdout. If a paper specified that all the segments from a single subject were assigned to only the training or the test set, we labeled that study as using ‘subject-based’ holdout. If a study used both segment-based and subject-based holdout in different analyses, we labeled the study as ‘both’. We labeled studies as ‘unclear’ if we could not determine whether the models were trained on segments of EEG recordings, and it was not explicitly stated that subjects were used as a factor in the holdout procedure.

## Results

### Data leakage leads to biased test-set accuracy

We analyze two datasets to test how the estimated accuracy of a DNN classifier depends on the train-test split. First, we examine the effects of data leakage in a patient-level classifier by training a model to diagnose Alzheimer’s disease. Second, we examine the effects of data leakage in a segment-level classifier by training a model to identify periods of time that include an epileptic seizure. In each of these analyses, we reuse a published DNN architecture to analyze an existing dataset.

### Identifying patients with Alzheimer’s disease

To determine whether segment-based holdout leads to a biased estimate of accuracy, we first trained a CNN to diagnose Alzheimer’s disease using segments of EEG. When the EEG segments were split into training and test sets without considering subject ID, the model showed nearly perfect test-set accuracy of 99.8% [99.1-100.0%] (Figure 1a). Performance quickly approached ceiling within the first 15 training epochs (Figure 2a). This high accuracy is consistent with prior studies that use segment-based holdout and report high accuracy for CNNs at identifying neurological disorders ^2,43,56^. Could this pattern of high accuracy reflect data leakage, instead of a robust and generalizable classifier?

**Figure 1:**
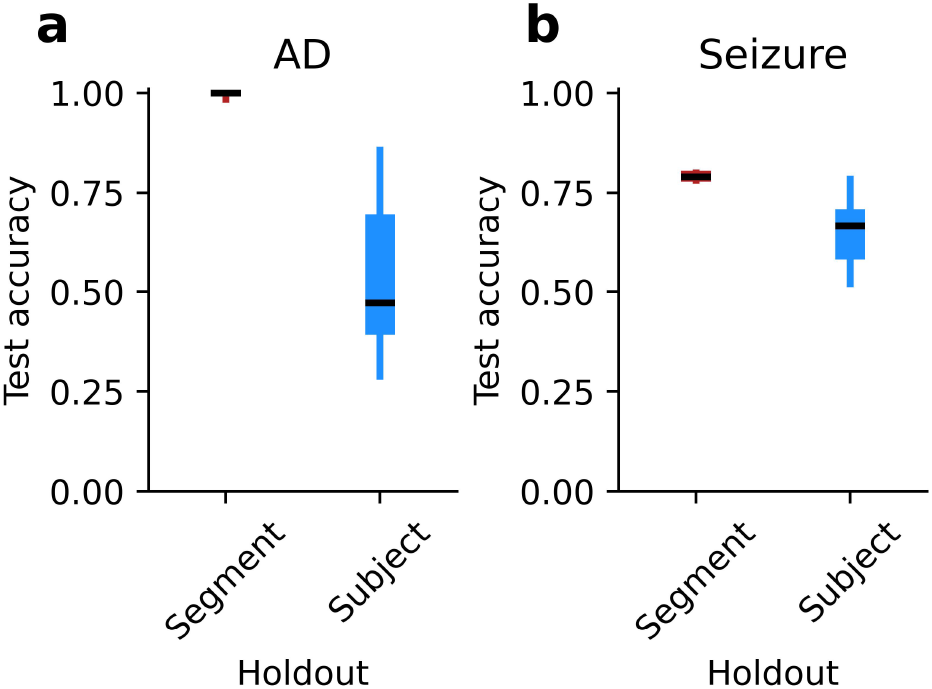
Test-set accuracy of CNN models predicting held-out data, plotted separately for segment-based holdout and subject-based holdout. (a) Accuracy for models trained to classify Alzheimer’s disease in individual subjects. Boxes show the inter-quartile range, dark lines show the median, and whiskers extend to the minimum and maximum points. (b) Accuracy for models trained to identify seizures in segments of EEG data. Details as in (a).

**Figure 2:**
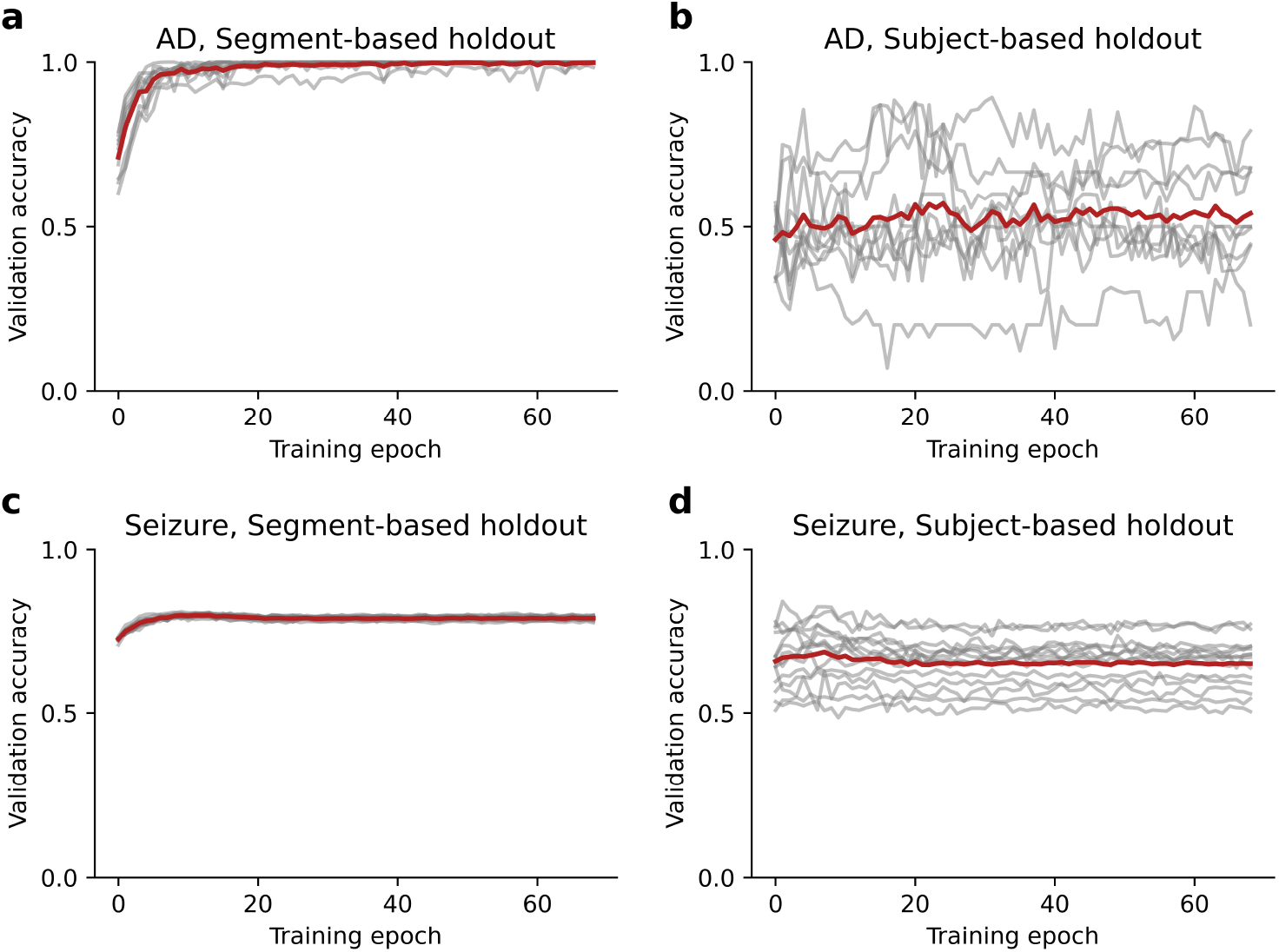
Test-set accuracy of CNN models plotted as a function of the training epoch. Grey lines show accuracy in individual cross-validation folds, and red lines show the average across folds. (a) Accuracy for models trained to classify Alzheimer’s disease using segment-based holdout. (b) Accuracy for models trained to classify Alzheimer’s disease using subject-based holdout. (c) Accuracy for models trained to identify seizures using segment-based holdout. (d) Accuracy for models trained to identify seizures using subject-based holdout.

When we used subject-based holdout, ensuring that individual subjects’ data did not appear in both the training and test sets, test accuracy dropped to 53.0% [43.1-64.8%], with 95% confidence intervals that included chance performance of 50%. Performance remained low throughout the training epochs (Figure 2b). Compared with subject-based holdout, segment-based holdout significantly overestimates the model performance on previously-unseen subjects (Wilcoxon *T* = 0.0, *p* = .002).

### Identifying segments containing epileptic seizures

In some cases, artificial neural network models have been used to identify time-limited events within ongoing brain activity, such as epileptic seizures. Does segment-based holdout also lead to data leakage when labeling periods of time within subjects? To answer this question, we trained a CNN to classify segments of EEG data as containing an epileptic seizure or not.

When the EEG segments were split into training and test sets without considering subject ID, the model reached a high test-set accuracy of 79.1% [78.8-79.4%] (Figure 1b). Accuracy leveled out within 10 training epochs (Figure 2c). When individual subjects’ data segments were restricted to appear in only the training or test set, however, accuracy fell to 65.1% [61.3-69.1%]. Accuracy remained low throughout training epochs (Figure 2d). Even when the model is tasked with labeling periods of activity within subjects, segment-based holdout significantly overestimates performance on previously-unseen subjects (Wilcoxon *T* = 0.0, *p* = 0.0001).

### Data leakage in published EEG studies

Do published translational EEG studies suffer from subject-specific data leakage, or do they avoid it by computing their test-set accuracy on held-out subjects? We examined the train-test split strategies in published studies that attempted to identify a clinical disorder using DNNs with EEG recordings. Out of the 63 relevant papers we found, only 17 (27.0%) unambiguously avoided this type of data leakage (Figure 3, Table 1). Leakage of subject-specific information is pervasive in the translational EEG literature.

**Figure 3:**
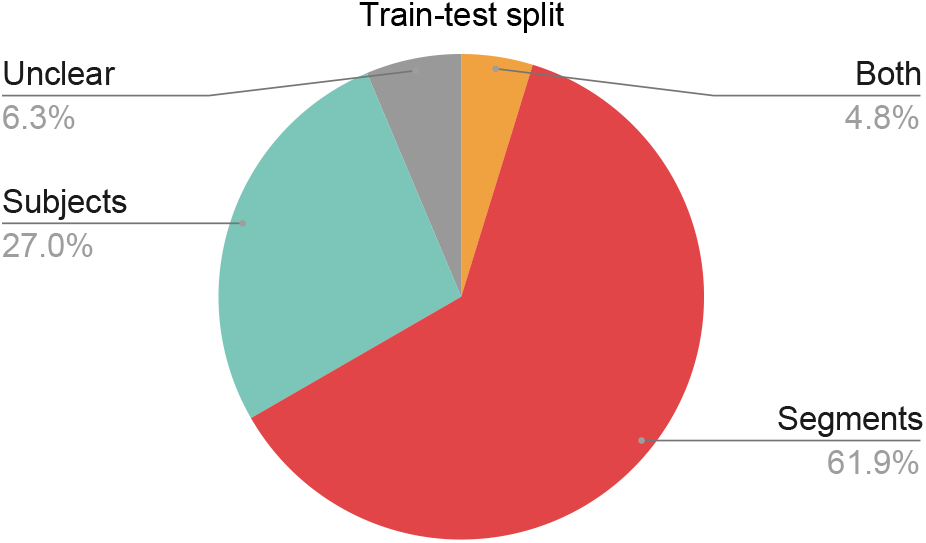
Number of studies using each type of test-split. “Segments”: Segments of EEG data were assigned to the training and test sets without regard to subject; this approach leads to data leakage. “Subjects”: Each subject’s data appeared in only the training set or the test set. “Both”: Both the Subjects and Segments approaches were used in different analyses. “Unclear”: We could not determine which approach was used for train-test splits.

**Table 1:**
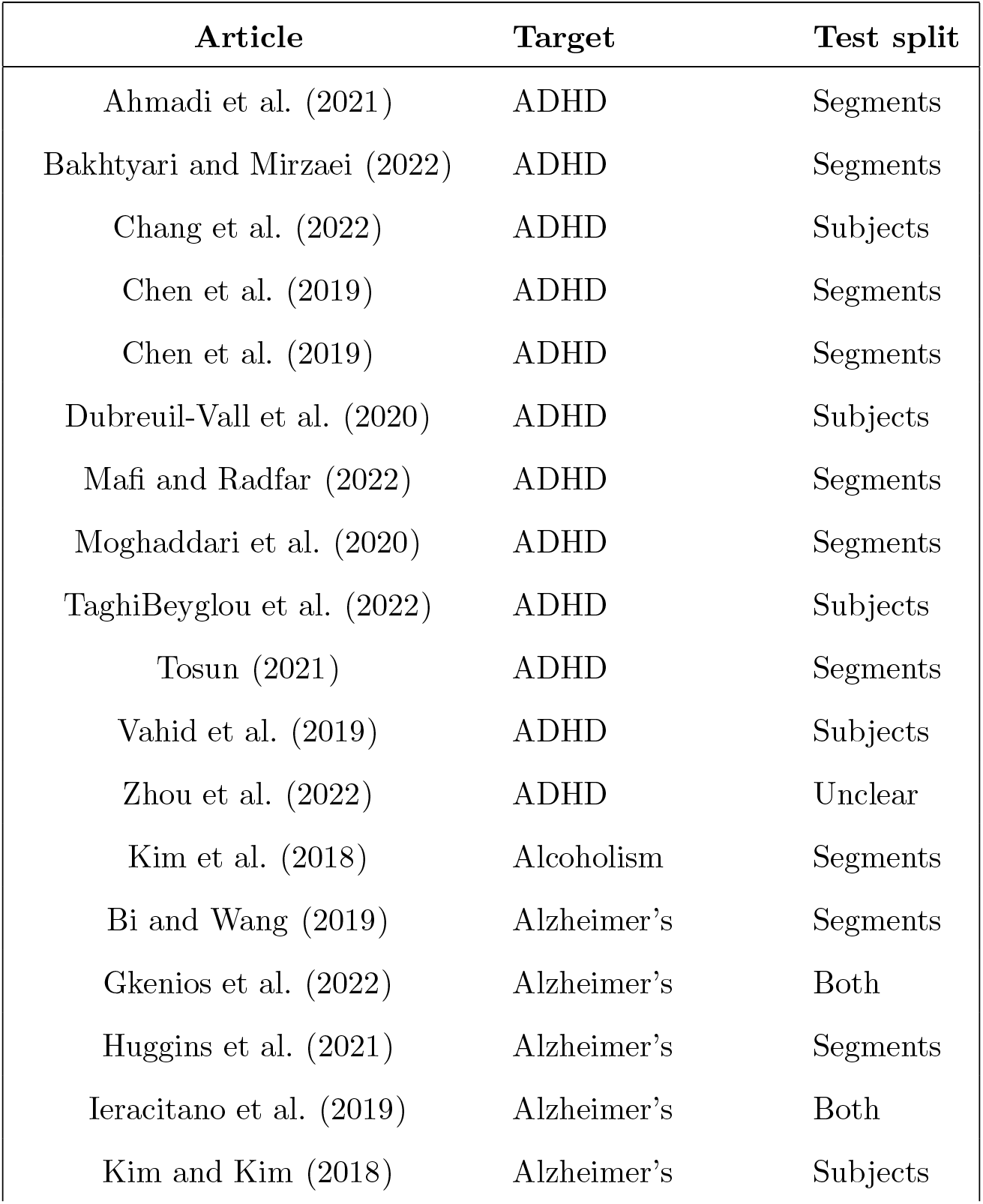
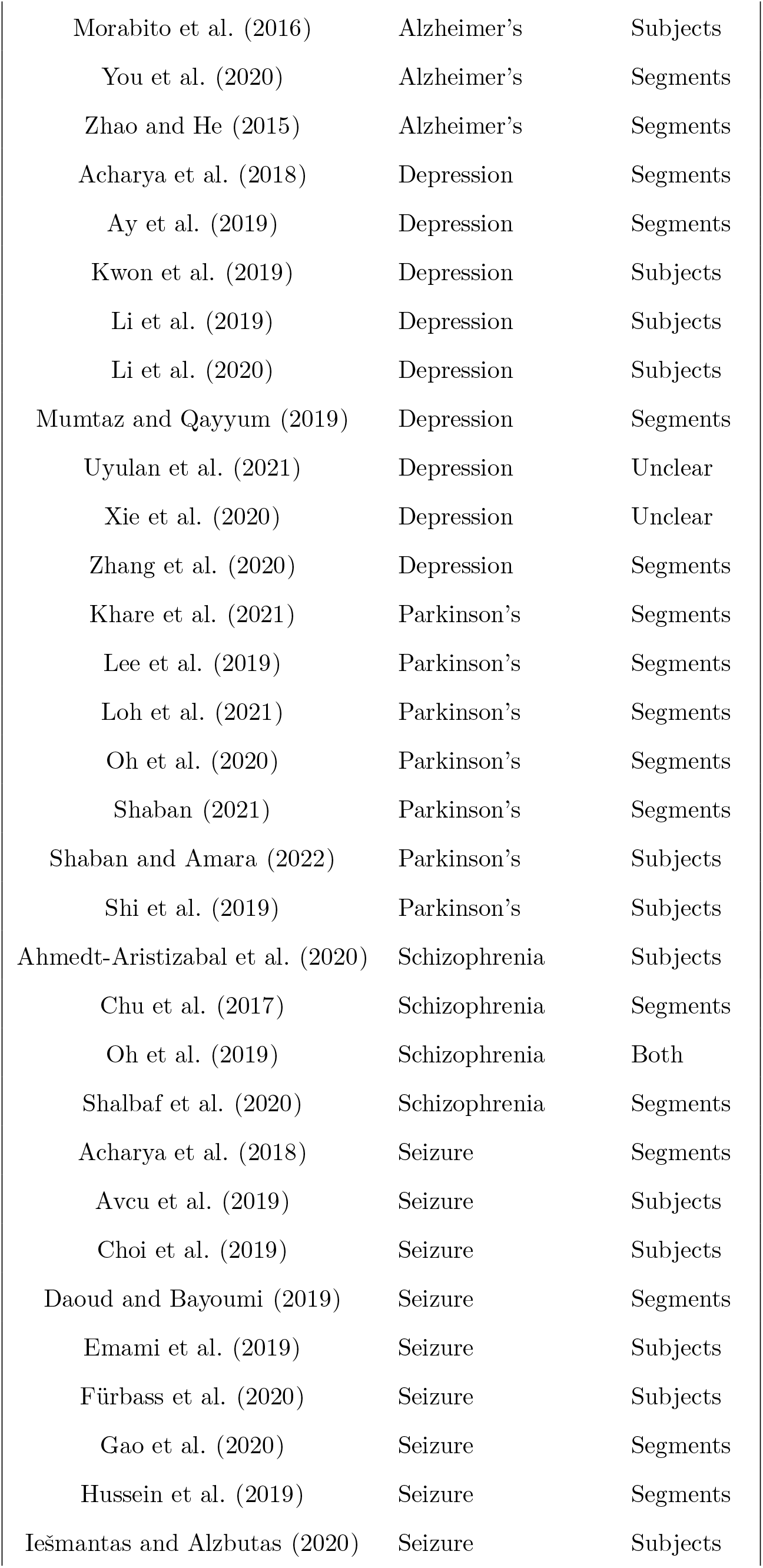
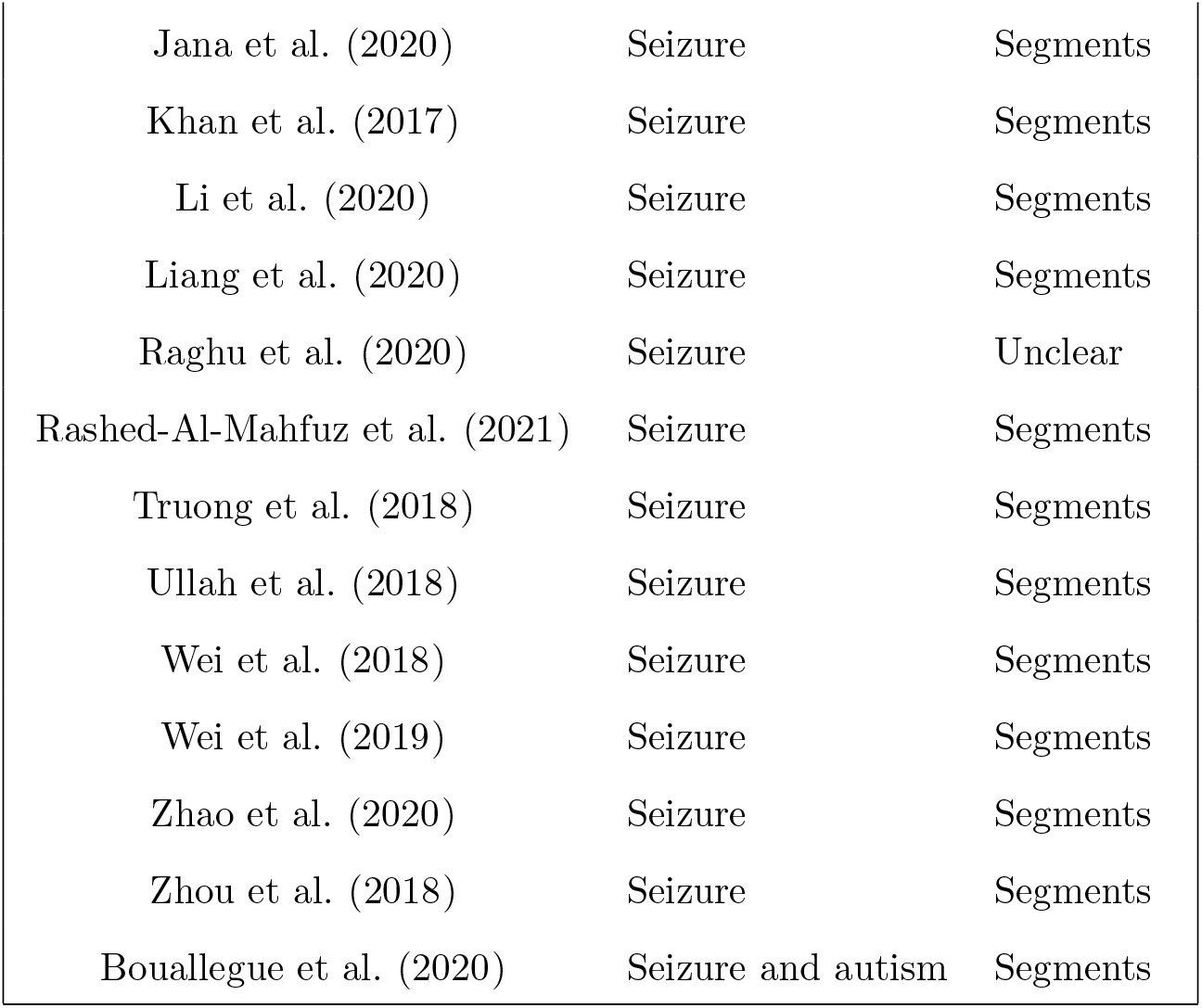
Prior translational studies using deep learning with EEG. Each line in the table describes one published translational study using a DNN with EEG data. The “Target” column holds the clinical condition being classified. The “Test split” column shows the approach used to determine how the data were divided into training and test sets. “Segments”: Segments of EEG data were assigned to the training and test sets without regard to subject; this approach leads to data leakage. “Subjects”: Each subject’s data appeared in only the training set or the test set. “Both”: Both the Subjects and Segments approaches were used in different analyses. “Unclear”: We could not determine which approach was used for train-test splits.

## Discussion

In EEG studies using deep learning, data leakage can occur when segments of data from the same subjects are included in both the training and test sets. Here we demonstrate that leakage of subject-specific information can dramatically overestimate the real-world clinical performance of a DNN classifier. Our Alzheimer’s CNN classifier appeared to have an accuracy of above 99% when using segment-based holdout, but its true performance on previously-unseen subjects was indistinguishable from chance. We found this bias in test-set performance both in a between-subjects task (identifying patients with Alzheimer’s disease) and in a within-subjects task (identifying segments that contain a seizure). Next, we show that this type of data leakage appears in the majority of published translational DNN-EEG studies we examined. Together, these results illustrate how an improperly-designed training-test split can bias the results of DNN studies, and show that biased results are widespread in the published literature.

To be useful in a clinical setting, a diagnostic classifier must be able to identify a disease in new patients. Models trained using segment-based holdout, however, strongly overestimate their ability to perform this task. Instead, these models may learn patterns associated with individual subjects, and then associate those idiosyncratic patterns with a diagnosis. As a consequence, performance of these models drops precipitously when they are tested in new subjects. When training a translational DNN classifier, the model must be tested with subjects who were not included in the training set.

### Data leakage when identifying events within subjects

Instead of identifying a disease in each subject, some studies attempt to identify a diseased process in each segment of time (see Appendix Table 1). DNN models of epilepsy, for example, often aim to classify the segments of data that contain a seizure. We demonstrated that those studies are not immune to data leakage in training-test splits: the accuracy in novel subjects is strongly overestimated when the test set includes subjects who were also in the training set. This result could arise if the model uses different patterns to identify seizures in each subject.

Subject-specific studies indicate that a bespoke classifier could be trained to identify seizures in each new patient ^34,46,47^. However, this would require every patient to have a large dataset of recordings that have already been labeled, which limits the clinical utility of this approach. A more realistic approach is to train DNN models to identify events in unseen patients.

### Data leakage in other methods

In studies which have only one observation per subject, cross-validation is trivial – single observations are simply assigned to the training or test set. However, in EEG and many other medical imagining methods, the data from each subject is routinely split into multiple segments. In this paper, we showed how data leakage can arise when a long recording is split into multiple shorter segments. However, the same principles apply to any other method that introduces statistical non-independence between the training and test sets. For example, some EEG-based DNNs treat every channel independently, and use information from each channel as a separate observation ^48^. Those studies are likely to suffer from substantial data leakage, since physiological sources of electrical activity appear redundantly across multiple EEG scalp electrodes ^50^.

These principles also apply to other medical imaging methods. Similar types of data leakage have been documented in studies using both functional ^73^ and anatomical ^74^ MRI, as well as in optical coherence tomography (OCT) ^65^.

## Conclusion

Data leakage occurs when EEG segments from one subject appear in the both the training and test sets. As a result, the test set accuracy dramatically overestimates the classifier’s performance in new subjects. This type of data leakage is common in published studies using DNNs and translational EEG. To accurately estimate a model’s performance, researchers must ensure that each subject’s data is included in only the training or the test set, but not both.

## Data Availability

EEG data for experiment 1 were provided by the Pacific Neuroscience Institute. These data are described by Ganapathi and colleagues (2022), and can be accessed through agreement with the authors of that study.
EEG data for experiment 2 were downloaded from the publicly-available Siena Scalp EEG Database (Detti et al, 2020a; 2020b) hosted on PhysioNet (Goldberger et al, 2000).
Paolo Detti. Siena Scalp EEG Database (version 1.0.0). PhysioNet, 2020a. URL https://doi.org/10.13026/5d4a-j060.
Paolo Detti, Giampaolo Vatti, and Garazi Zabalo Manrique de Lara. EEG synchronization analysis for seizure prediction: A study on data of noninvasive recordings. Processes, 8(7):846, 2020b.
Aarthi S Ganapathi, Ryan M Glatt, Tess H Bookheimer, Emily S Popa, Morgan L Ingemanson, Casey J Richards, John F Hodes, Kyron P Pierce, Colby B Slyapich, Fatima Iqbal, et al. Differentiation of subjective cognitive decline, mild cognitive impairment, and dementia using qEEG/ERP-based cognitive testing and volumetric MRI in an outpatient specialty memory clinic. Journal of Alzheimer's Disease, pages 1-9, 2022 (preprint)
Ary L Goldberger, Luis AN Amaral, Leon Glass, Jeffrey M Hausdorff, Plamen Ch Ivanov, Roger G Mark, Joseph E Mietus, George B Moody, Chung-Kang Peng, and H Eugene Stanley. PhysioBank, PhysioToolkit, and PhysioNet: Components of a new research resource for complex physiologic signals. Circulation, 101(23):e215-e220, 2000.

https://physionet.org/content/siena-scalp-eeg/1.0.0/

## Acknowledgements

This work was supported by SPARK Neuro, Inc.

## Appendix

### AD model definition

**Listing 1:**
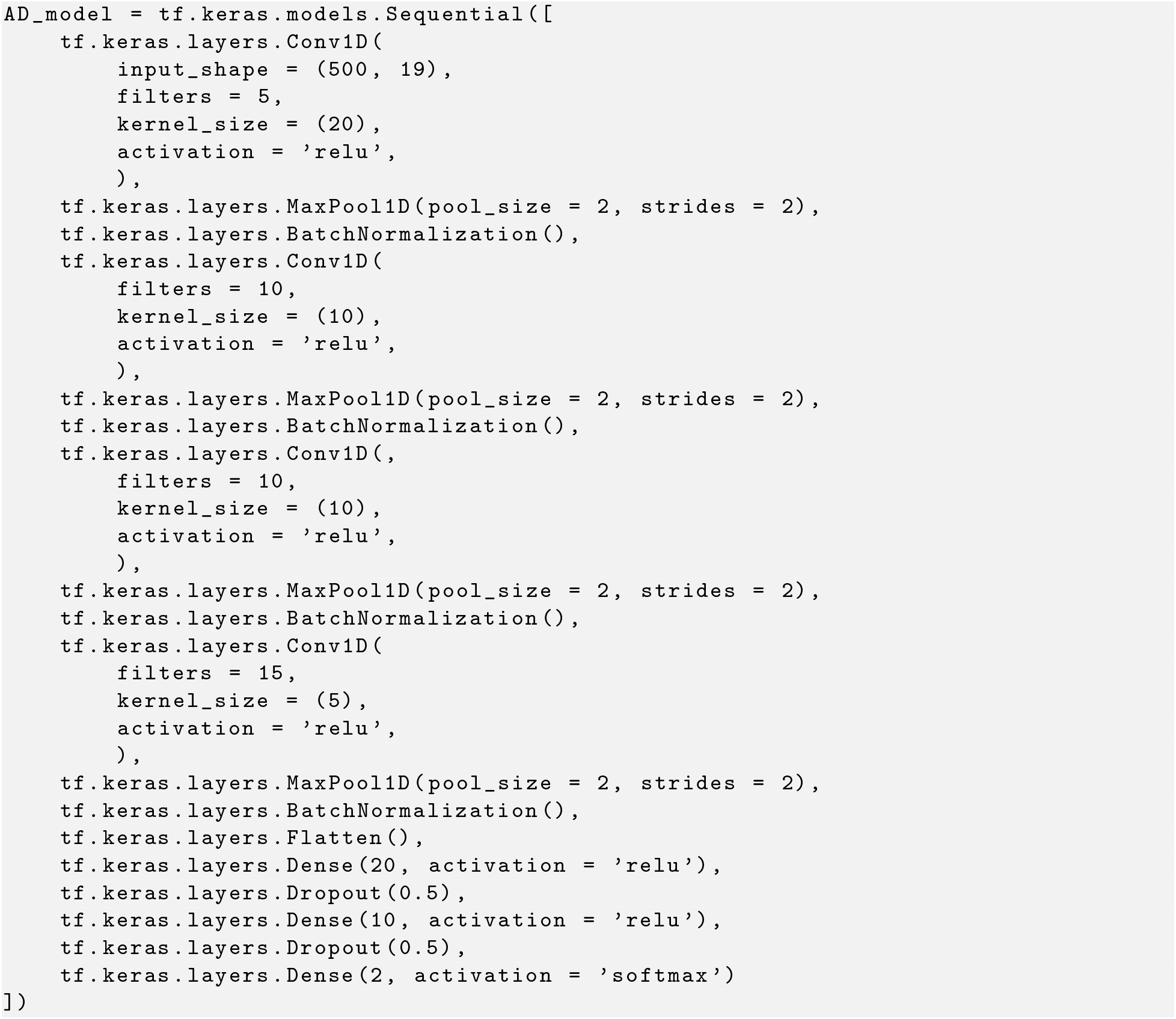
AD model definition

### Seizure model definition

**Listing 2:**
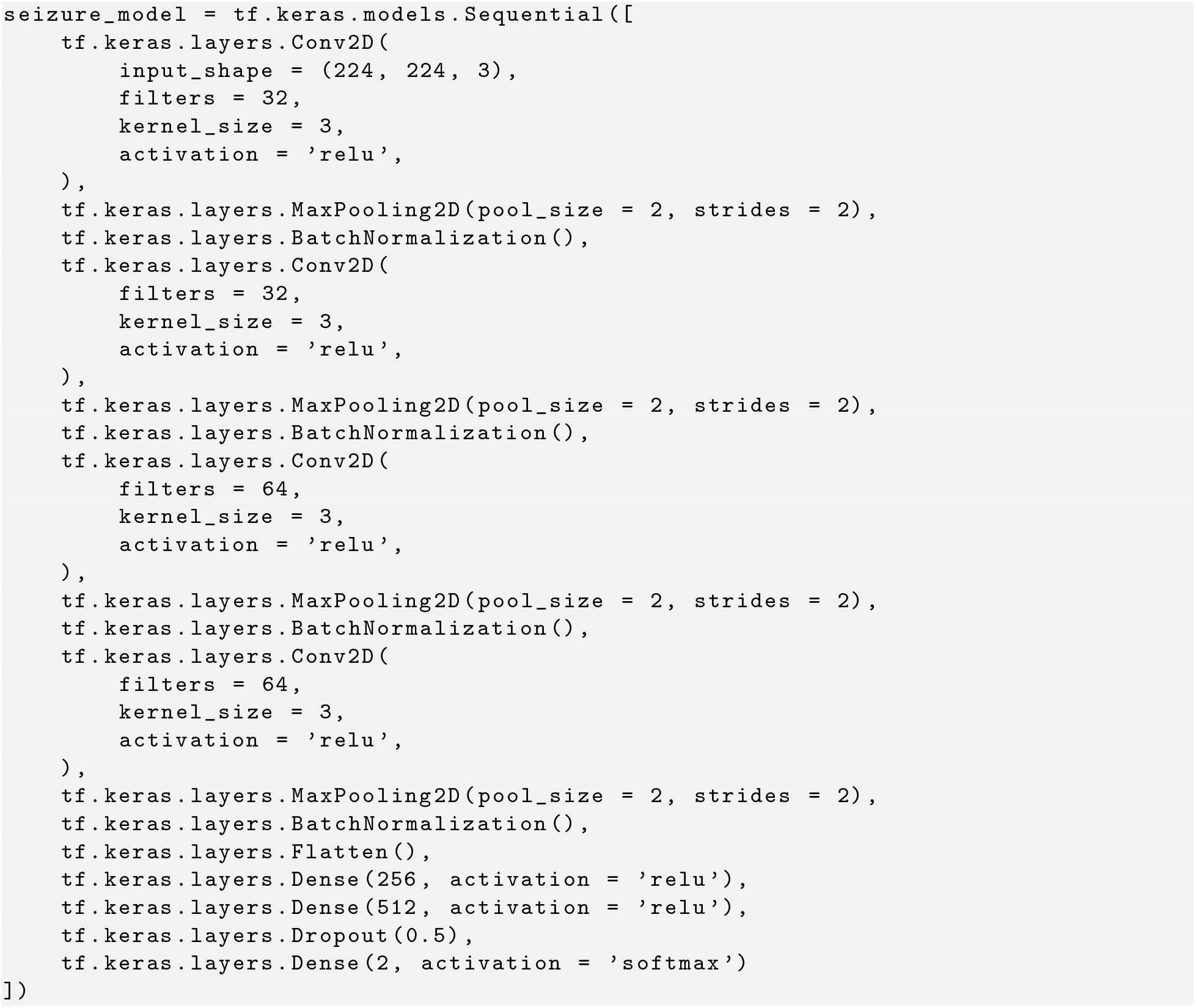
Seizure model definition

